# Quantification of SARS-CoV-2 and cross-assembly phage (crAssphage) from wastewater to monitor coronavirus transmission within communities

**DOI:** 10.1101/2020.05.21.20109181

**Authors:** Hyatt Green, Maxwell Wilder, Mary Collins, Ariana Fenty, Karen Gentile, Brittany L. Kmush, Teng Zeng, Frank A. Middleton, David A. Larsen

**Affiliations:** Department of Environmental and Forest Biology, SUNY-ESF, Syracuse, NY 13210; Department of Environmental Studies, SUNY-ESF, Syracuse, NY 13210; Department of Neuroscience and Physiology, Upstate Medical University, Syracuse, NY 13210; Department of Public Health, Syracuse University, Syracuse, NY 13244; Department of Civil and Environmental Engineering, Syracuse University, Syracuse, NY 13244

## Abstract

Wastewater surveillance of SARS-CoV-2 has become an attractive tool for combating the spread of COVID-19 by assessing the presence or levels of the virus shed in a population. However, the methods to quantify viral RNA and to link those quantities to the level of infection within the community vary. In this study, we sought to identify and optimize scalable methods for recovery of viral nucleic acids from wastewater and attempted to use a constitutive member of the gut virome, human-specific crAssphage, to help account for unknown levels of SARS-CoV-2 decay and dilution in the wastewater infrastructure. Results suggest that ultracentrifugation of a small volume of wastewater through a 50% sucrose cushion followed by total nucleic acid extraction yielded quantifiable virus in an area with a modest number of COVID-19 cases. Further, the ratio of log_10_(SARS-CoV-2):log_10_(crAssphage) appears to be associated with the cumulative incidence of COVID-19 in the Syracuse, NY area. In areas where ultracentrifuges are available, these methods may be used to link SARS-CoV-2 quantities in wastewater to levels of transmission within communities with sewer service.

## Introduction

While the primary mechanism of transmission of Sudden Acute Respiratory Syndrome Coronavirus 2 (SARS-CoV-2), the virus that causes the respiratory disease Coronavirus Disease 2019 (COVID-19), is through respiratory droplets, a significant number of inactive viral particles are shed in the feces of infected persons^1–3^. At the very least, the prospect of surveilling possible disease burden via surveillance of untreated wastewater offers opportunities for monitoring the emergence of SARS-CoV-2 transmission^4^ and trends over time^5^ including any eventual reduction in transmission. When coupled with information from standard surveillance methodology, such a surveillance system could conceivably and efficiently inform decisions about where to focus resources (e.g., individual swab testing, contact tracing), where to target interventions such as social distancing, as well as how and at what rate to reduce broad scale social distancing and reopen local economies^6^. The tool could also be used to efficiently monitor potentially vulnerable facilities, such as jails, schools, or assisted-living facilities.

Methods to recover SARS-CoV-2 from wastewater have varied widely in the few recent reports with implications for their cost, scalability, and susceptibility to disruptions in supply chains. Medema and colleagues used moderate speed centrifugation methods with centrifuge filters (Centricon® Filters) to successfully concentrate and purify viral particles/genomes^4^. This method was somewhat successfully replicated by another group that also had some success by filtration through electronegative membranes^7^. However, the cost and availability of centrifuge filters can be limiting for some laboratories, especially considering to likelihood of supply chain disruptions during a pandemic. Perhaps the most scalable method that requires very little specialized equipment was proposed by Wu and colleagues where 0.2 micron filtrates were PEG-precipitated and then Trizol™ extracted to obtain purified RNA^8^. Among the simplest procedures reported, assuming the equipment is available, is the ultracentrifugation of small volumes (11 ml) of wastewater for one hour followed by extracting nucleic acids from the pellet^5^. Aluminum-driven flocculation followed by nucleic acid extraction has also been used to successfully detect the virus in wastewater^9^. Tangential flow filtration followed by PEG-precipitation has also been used to concentrate and purify viruses from wastewater, but might pose problems in scaling-up due to the length of time needed to filter a single sample^10^. In the anticipation that wastewater surveillance for SARS-CoV-2 and other pathogens will become widespread, methods that are sensitive, scalable, and cost-effective will be needed by regional and local laboratories.

Methodological differences aside, the question of how SARS-CoV-2 concentrations in wastewater can have a significant bearing on public health decisions remains. An attractive prospect is to use viral concentrations in wastewater to estimate the number of infected individuals “upstream” in the represented catchment area. Pairing results from wastewater with shedding rates to arrive at the estimated number of infected individuals in the represented area provided a higher than expected number of cases in the Boston area^8^ and highly uncertain estimates in Southeast Queensland^7^. Factors that remain problematic to this approach and others are the low concentrations of virus obtained, which thereby increases the uncertainty in the quantification process, as well as issues regarding viral fate and transport throughout the wastewater infrastructure.

In this study, our goals were to: a) detect and, if possible, quantify SARS-CoV-2 in wastewaters of the upstate New York area (Onondaga County, NY, US), b) improve scalable methods of wastewater sample processing to maximize the recovery of viral nucleic acids, and c) integrate the measurements of both SARS-CoV-2 and an abundant gut-derived bacteriophage (crAssphage) which may help account for decay and dilution in wastewater infrastructure. Since we also sought an inexpensive, effective method that could be brought to scale, ultracentrifugation seemed attractive because few supplies were needed, which allowed us to bypass some supply chain issues, and many research institutions in the region have the required equipment on site. Our results indicate that ultracentrifugation incorporating a 50% sucrose cushion was successful at concentrating viral nucleic acids while removing impurities. As a result, we were able to detect, and in most cases, quantify SARS-CoV-2 in small volumes (20 ml) of wastewater collected from an area with a moderate number of COVID-19 cases. We also show that the ratio of SARS-CoV-2:crAssphage visually correlates with the cumulative incidence of COVID-19.

## Methods

### Wastewater Ultracentrifugation

Twenty-four hour composite wastewater samples (1.9 L) were collected from 11 access points (i.e., wastewater treatment plants, influent pump stations, or interceptor lines) in Syracuse, NY and other locations in Onondaga County, NY on May 6th and 13th, 2020 (Table 1). Samples were stored at 4°C following collection and transported on ice to Upstate Medical University (Syracuse, NY) for processing the next morning. Upon receipt, samples were mixed to resuspend particulates and 20 ml was aliquoted to a 38.5 ml ultracentrifuge tube (ThermoFisher, #750000471). A 12 ml sucrose cushion (50% sucrose in TNE buffer [20 mM Tris-HCl (pH 7.0), 100 mM NaCl, 2 mM EDTA]) was carefully added underneath the wastewater with a 10 ml disposable serological pipette so that two distinct layers were formed (Figure 1A). Fifty percent sucrose yielded higher crAssphage DNA concentrations than 20% or 70% (Table S1). In batches of six, samples were then purified by centrifugation at 150,000 x g at 4°C for either 90 minutes (May 6) or 45 minutes (May 13) on a Sorvall WX Ultra series with a Sorvall Surespin™ 630 rotor (ThermoFisher). Centrifugation times of 45 and 90 minutes provided roughly the same recovery of viral nucleic acids (Table S2). Supernatant was then decanted, and pellets (Figure 1B) were resuspended in 200 μL 1X PBS. Pellets were stored at −20°C for <12 hours prior to nucleic acid extraction. Distilled water (20 ml) was used as a processing blank.

**Table 1.**
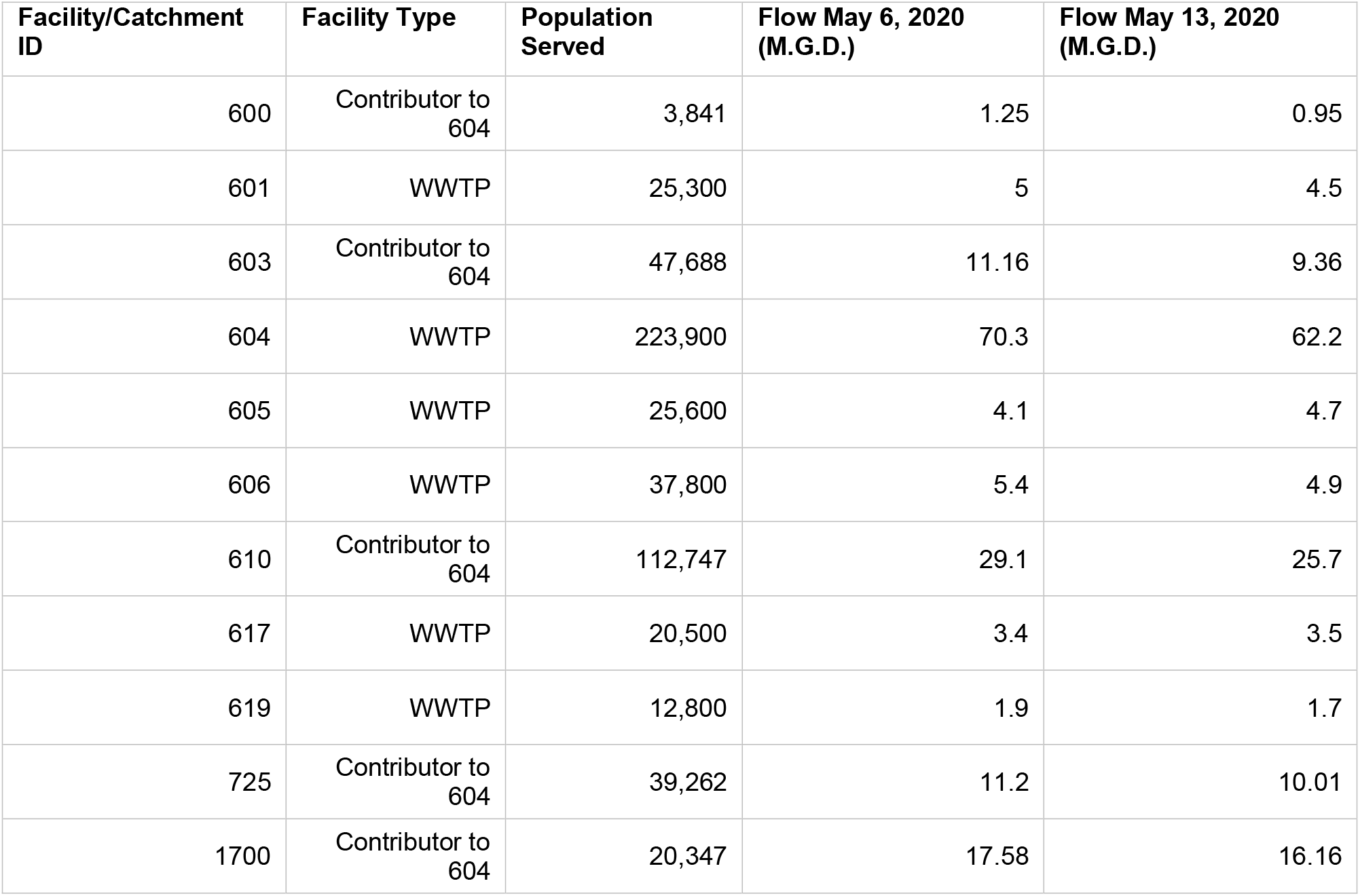
Facility characteristics and average flow (millions of gallons per day) on May 6 and 13, 2020.

**Figure 1.**
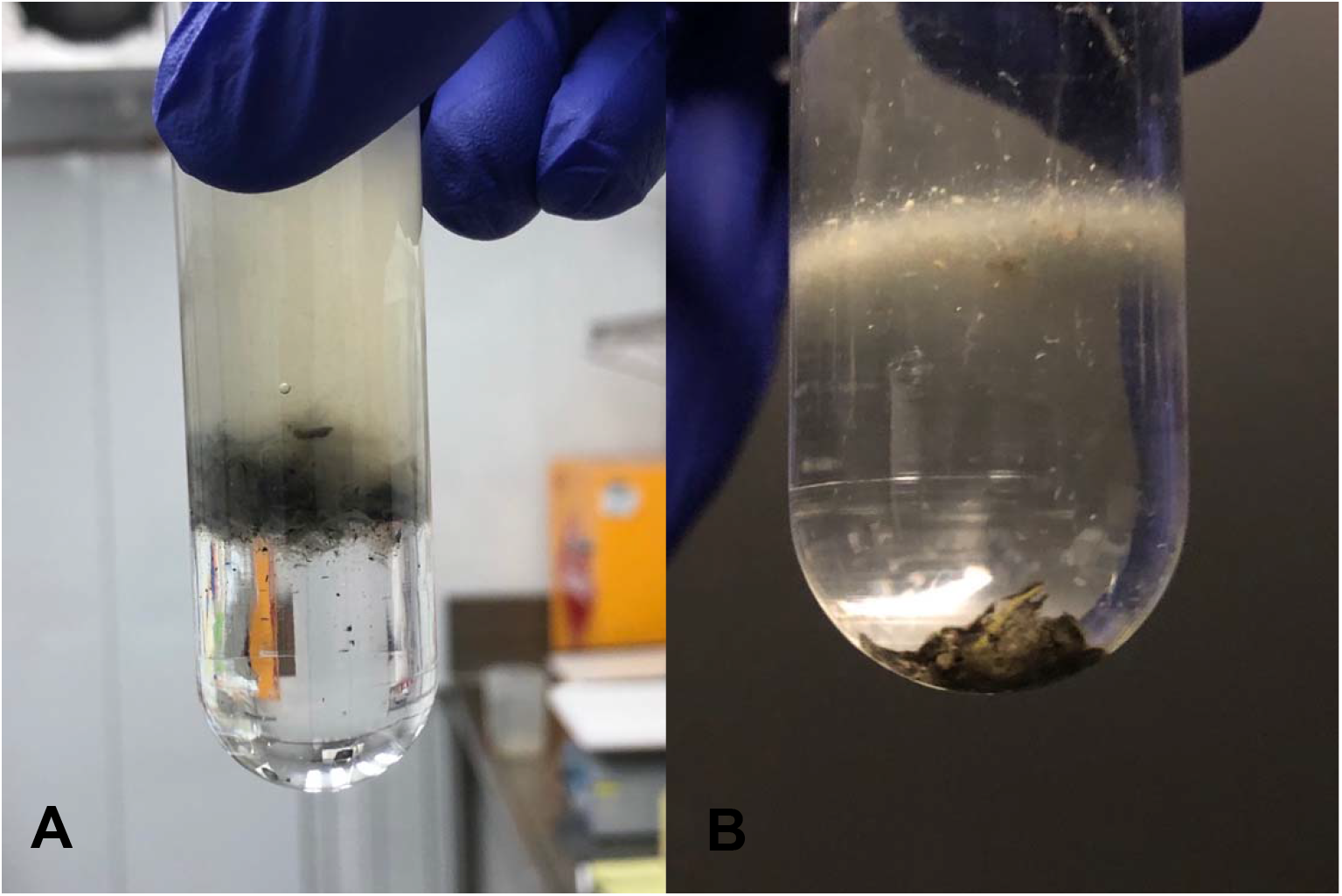
(**A**) Raw wastewater above a 50% sucrose solution prior to ultracentrifugation. (**B**) Pellet produced by ultracentrifugation and residual debris on top of the 50% sucrose cushion.

### Nucleic Extraction and Synthesis of crAssphage cDNA

Total nucleic acids were extracted using the AllPrep® PowerViral® DNA/RNA Kit (Qiagen, Hilden, Germany) according to manufacturer’s protocol. Samples were eluted in 50 μL RNase free water. Extraction blanks using distilled water were performed in each extraction batch to assess contamination. To assess the recovery of crAssphage RNA through the methods previously described, cDNA was generated using the QuantiTect Reverse Transcription Kit (Qiagen) according to manufacturer’s protocol.

### CrAssphage qPCR

qPCR was used to detect the presence of crAssphage using the previously developed CPQ_056 assay^11^ (Table 2). Reactions consisted of 12.5 μL TaqMan® Environmental MasterMix (ThermoFisher), 1 μM primers, 80 nM probe, molecular grade water, and 2 μL template DNA for a total reaction volume of 25 μL. Each reaction was run on either a QuantStudio3™ or QuantStudio5™ (ThermoFisher) under the following thermal cycling conditions: 10 minutes at 95°C, followed by 40 cycles of 95°C for 15 seconds and 60°C for 1 minute. A DNA standard was generated by purification of amplified PCR product using Roche High Pure PCR Template Preparation Kit. Standard DNA quantity was assessed on NanoDrop spectrophotometer and Qubit® fluorometer. A standard curve, consisting of purified amplicons ranging from 1 × 10^6^ to 5 copies/reaction, was used to convert Ct values to gene copies per reaction (Table 3). A CPQ_056 assay modified for SYBR Green chemistry was used in some optimization trials (see Supplementary).

**Table 2.**
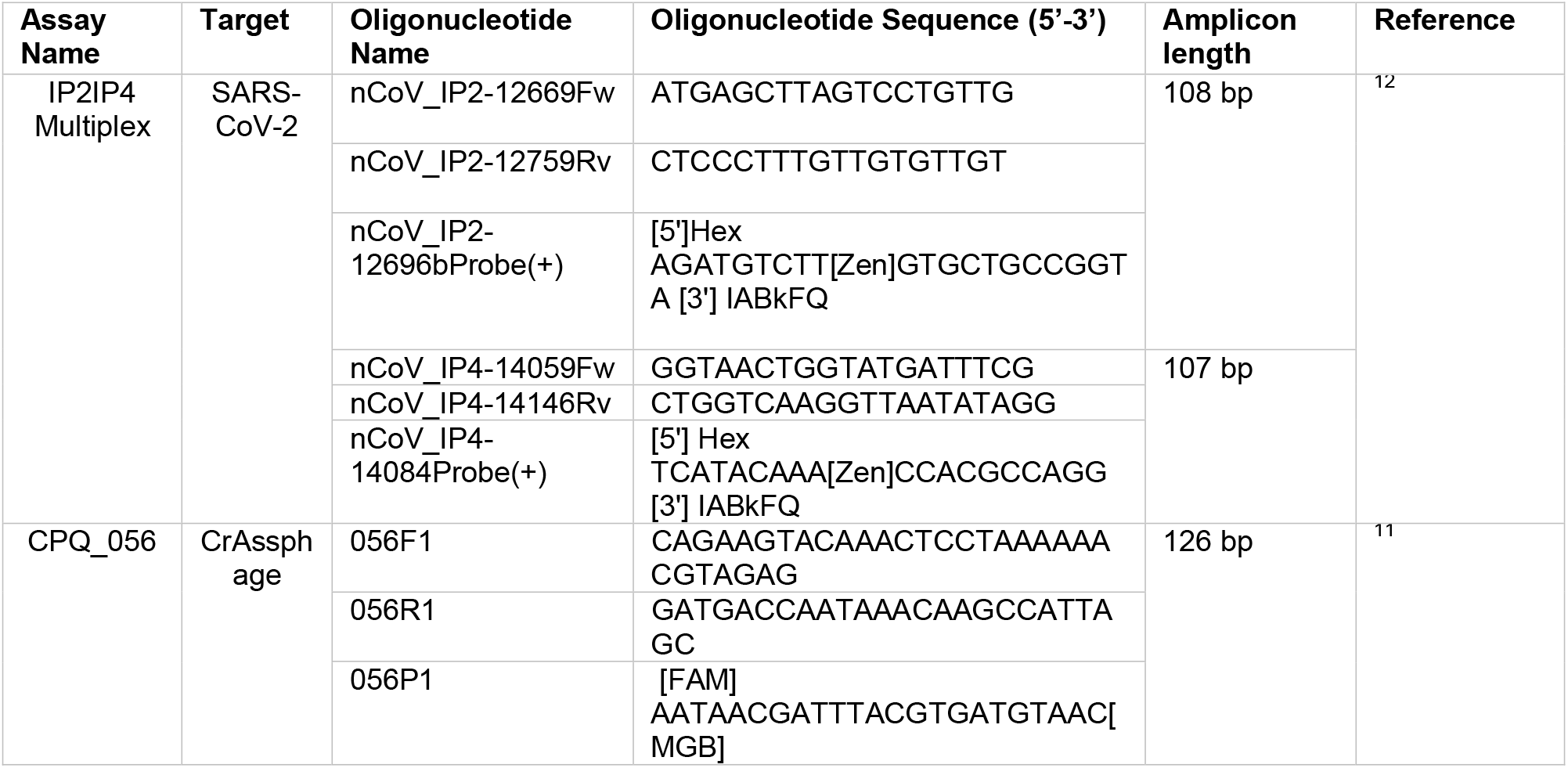
qPCR assays used.

**Table 3.** qPCR assay performance parameters.

| Assay | R<sup>2</sup> | Intercept | Slope | Efficiency |
| --- | --- | --- | --- | --- |
| CPQ_056 | 0.974 | 39.568 | -3.406 | 0.97 |
| IP2IP4 | 0.849 | 41.026 | -3.508 | 0.93 |

### SARS-CoV-2 RT-qPCR

To detect SARS-CoV-2, RT-qPCR was used with a multiplex reaction containing the previously published IP2 and IP4^12^ assays (Table 2). Reactions consisted of 6.25 uL Reliance One-Step Multiplex RT-qPCR Supermix, 0.4 μM each primer, 0.16 μM probes, molecular grade water, and 2.5 uL template total nucleic acids for a total reaction volume of 25 μL. Thermal cycling conditions were 10 minutes at 50°C, 10 minutes at 95°C, followed by 45 cycles of 95°C for 10 seconds and 59°C for 30 seconds. A standard curve, consisting of amplicons ranging from 250 to 2.5 copies/reaction, was used to convert Ct values to gene copies per reaction (Table 3). All no-template controls for both crAssphage and IP2IP4 (n > 40) were negative throughout the entire study.

### Recovery of crAssphage and SARS-CoV-2

To assess the proportion of viral nucleic acids remaining in each phase (including the pellet) after centrifugation, wastewater samples (2 x 20 ml) were spiked with SARS-CoV-2 (resulting in approximately 580 gene copies per ml wastewater). Pellets were generated by ultracentrifugation at 150,000 x g for 45 minutes and the following layers were removed; aqueous upper (top 10 ml), aqueous lower (second 9 ml), cushion interface (1.5 ml, specifically targeting visible suspended particles on top of sucrose layer), sucrose upper (6 ml), sucrose lower (6 ml), and pellet (appx. 200 μL). A 200 aliquot of each layer was extracted using the PowerViral kit (Qiagen) and extracts were analyzed with the IP2IP4 assay in 25 μL reaction volumes under the conditions described previously (Table S6).

### Cumulative Incidence of COVID-19

To correlate evidence of SARS-CoV-2 transmission in wastewater with cases of COVID-19 in the wastewater catchment area we retrieved the total number of COVID-19 cases by ZIP code from the Upstate Hospital electronic medical record system. These records reflect approximately 40% of the total COVID-19 cases in Onondaga County. We standardized the number of COVID-19 cases by ZIP code to the ZIP code population to calculate a cumulative incidence per 1,000 residents. We then visualized cumulative incidence alongside the ratio of log_10_(SARS-CoV-2):log_10_(crAssphage).

## Results

### Recovery of Viral Nucleic Acids

Direct ultracentrifugation of a wastewater sample through a %50 sucrose cushion resulted in the formation of a translucent, but visible, pellet often bordered by darker, lower density residue, presumably organics, metal sulfides, and/or other impurities in the wastewater (Figure 1). Depending on the wastewater sample, lower density impurities were often resting at the cushion interface. Recovery trials with inactive SARS-CoV-2 spiked samples indicated that no quantifiable SARS-CoV-2 RNA, crAssphage RNA, or crAssphage DNA remained in the upper aqueous phase or sucrose layer after centrifugation (Table 4). Given that only a portion of each layer was tested, it’s possible there may have been some non-pelleted residual viral nucleic acids at concentrations below the limit of detection, but it is clear that the vast majority of SARS-Cov-2 and crAssphage viral nucleic acids are pelleted under these conditions. After nucleic acid extraction and qPCR, we estimated recovery of SARS-CoV-2 based on the addition of a known quantity of SARS-CoV-2 to be about 12% attributing some loss to non-pelleted viral RNA, but most to the subsequent nucleic acid extraction procedure.

**Table 4.** Recovery of SARS-CoV-2 RNA through a sucrose cushion.

| Layer | Replicate Tube | Mean Copies SARS-CoV-2<br>(+/- SD) | SARS-CoV-2<br>Rxns Positive<br>(out of three) |
| --- | --- | --- | --- |
| Aqueous Upper | 1 | <LOQ | 0 |
|  | 2 | <LOQ | 0 |
| Aqueous Lower | 1 | <LOQ | 0 |
|  | 2 | <LOQ | 0 |
| Cushion Interface | 1 | <LOQ | 0 |
|  | 2 | <LOQ | 2 |
| Sucrose Upper | 1 | <LOQ | 0 |
|  | 2 | <LOQ | 0 |
| Sucrose Lower | 1 | <LOQ | 0 |
|  | 2 | <LOQ | 0 |
| Pellet | 1 | 1.37E3 (7.39E2) per pellet | 3 |
|  | 2 | 1.42E3 (7.19E2) per pellet | 3 |

### Yield and Quality Assessment of Total RNA

The direct ultracentrifugation and purification of wastewater resulted in recovery of a considerable amount of total RNA. In the samples that were collected on May 6 the average yield was 26.3 ng/μL (std. dev. = 11.7) (Figure S2, top panel). However, these estimates were likely affected by the presence of considerable DNA carryover. Thus, for the May 13 samples, the addition of DNase produced lower yields, with an average of 2.5 ng/μL (std. dev. = 2.3) (Figure S2, bottom panel). The RNA integrity numbers of the samples were not significantly different in either batch, however, with May 6 average of 4.3 ng/μL (std. dev. = 0.8) and a May 13 average of 3.4 ng/μL (std. dev. = 1.9) (Figure S2). Notably, these RNA integrity values are similar to ones obtained from human biofluid waste products, such as saliva, urine, or fecal matter.

### Detection and Quantification of Viral Nucleic Acids in Wastewater

Over a two-week period, we detected some level of SARS-CoV-2 in 18 out of 22 samples, 13 of which were in the quantifiable range (Table 5). SARS-CoV-2 was more prevalent in the May 13 samples (detected in 11 out of 11 samples) compared to the May 6 samples (7 out of 11), possibly due to a significantly lower flow on May 13 (paired t-test, *p*=0.045). If any of the three reaction wells crossed the fluorescence threshold, the sample was interpreted as positive due to all negative controls throughout the study testing negative. Likewise, SARS-CoV-2 fell within the quantifiable range in 9 out of 11 May 13 samples and only 4 out of 11 May 6 samples. The average number of SARS-CoV-2 genome copies within quantifiable samples over the two week period was 42.7 (std.dev = 32.9) genomes/ml while the highest observed was 112.35 (std. dev. = 8.01) genome/ml of wastewater.

**Table 5.** Concentrations of SARS-CoV-2 and crAssphage DNA and RNA in sampled catchments.

| Date Collected | Facility/<br>Catchment | Mean copies crAssphage<br>(CPQ_056)/mL wastewater (+/- SD) |  | SARS-CoV-2 |  |
| --- | --- | --- | --- | --- | --- |
|  |  | DNA | RNA | Mean Copies<br>SARS-CoV-2 /<br>mL<br>(+/- SD) | Rxns Positive<br>(out of three) |
| May 6 | 600 | 5.79E4 (3.58E3) | 2.76E2 (4.50E0) | <LOQ | 0 |
| May 6 | 601 | 1.76E5 (3.36E4) | 5.33E2 (1.33E1) | 1.12E2 (8.01E0) | 3 |
| May 6 | 603 | 5.44E4 (4.03E3) | 1.65E2 (3.75E1) | 1.51E1 (7.89E0) | 3 |
| May 6 | 604 | 9.29E4 (5.65E3) | 3.95E2 (5.08E1) | 4.59E1 (1.29E1) | 3 |
| May 6 | 605 | 1.19E5 (1.26E4) | 3.31E2 (7.55E1) | <LOQ | 0 |
| May 6 | 606 | 7.36E4 (1.47E3) | 2.48E2 (4.61E1) | <LOQ | 2 |
| May 6 | 610 | 3.33E4 (1.19E3) | 1.37E2 (1.59E1) | 2.25E1 (1.45E1) | 3 |
| May 6 | 617 | 1.47E5 (8.78E3) | 2.73E2 (5.12E1) | <LOQ | 2 |
| May 6 | 619 | 7.58E4 (1.18E4) | 1.62E2 (2.35E1) | <LOQ | 0 |
| May 6 | 725 | 1.01E5 (8.28E3) | 2.32E2 (2.99E1) | <LOQ | 0 |
| May 6 | 1700 | 2.50E4 (2.25E3) | 1.02E2 (2.30E1) | <LOQ | 1 |
| May 13 | 600 | 1.07E5 (6.97E3) | 1.03E2(2.44E1) | 7.50E0(2.94E0) | 3 |
| May 13 | 601 | 8.44E4 (7.8E33) | 1.40E2 (3.44E1) | 2.82E1 (4.67E0) | 3 |
| May 13 | 603 | 6.43E4 (1.43E3) | 1.32E1 (6.52E0) | 3.04E1 (3.14E1) | 3 |
| May 13 | 604 | 6.74E4 (1.03E3) | 5.50E1 (1.57E1) | 5.72E1 (2.62E1) | 3 |
| May 13 | 605 | 9.15E4 (1.81E3) | 3.94E1 (1.57E1) | 1.20E1 (2.39E0) | 3 |
| May 13 | 606 | 9.65E4 (1.38E3) | 4.00E1 (1.60E1) | 4.90E1 (6.33E0) | 3 |
| May 13 | 610 | 2.21E4 (1.02E3) | <LOQ (0/3) | 6.92E1 (7.89E0) | 3 |
| May 13 | 617 | 9.81E4 (3.15E3) | 4.90E1 (1.17E1) | <LOQ | 1 |
| May 13 | 619 | 1.54E5 (7.35E3) | 4.21E1 (8.88E0) | <LOQ | 1 |
| May 13 | 725 | 1.95E5 (7.73E3) | 1.13E2 (7.16E0) | 1.30E1 (8.34E0) | 3 |
| May 13 | 1700 | 4.96E5 (3.66E3) | 8.29E1 (3.43E1) | 9.41E1 (2.00E1) | 3 |

In contrast, crAssphage DNA was abundant in every sample analyzed with an average of 1.11 x 10^5^ copies/ml across the two week period with no significant difference between the two sample sets. crAssphage RNA was detected in every sample except the sample from Facility 610 on May 13. crAssphage RNA was much less abundant than crAssphage DNA with an average of 1.68 x 10^2^ copies/ml. Interestingly, while there was no significant difference in crAssphage DNA between the two sample sets, crAssphage RNA was significantly lower on May 13 than May 6 (paired and unpaired t-tests, *p* < 0.001).

### Association Between crAssphage Loads and Population Served

While crAssphage DNA concentrations were not significantly associated with population served, flow, SARS-CoV-2 concentrations, or crAssphage RNA concentrations, there was a significant linear relationship between the loads of both crAssphage DNA (*p* < 0.001) and RNA (*p* < 0.01) and population served in each catchment (Figure 2).

**Figure 2.**
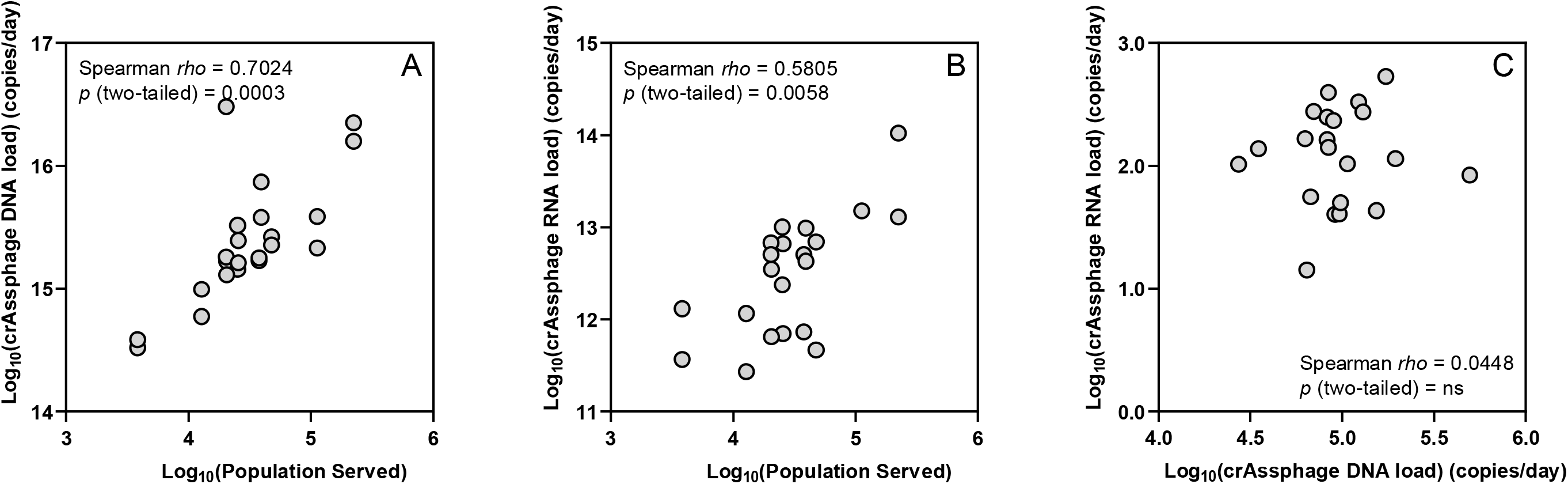
Relationship between crAssphage DNA load and population served (A), crAssphage RNA load and population served (B), crAssphage RNA load and crAssphage DNA load (C). Loads are simply calculated as the concentration x the flow rate.

### Spatial Association Between SARS-CoV-2:crAssphage Ratios in Wastewater and COVID-19 Incidence

Although the number of cases in each catchment would allow a better assessment of the relationship between viral concentrations in wastewater and the level of transmission in the respective community, visual inspection suggests a spatial correlation between the cumulative incidence of cases by zip code from the Upstate hospital system and the ratio of SARS-CoV-2:crAssphage in wastewater with higher ratios occurring in areas of higher incidence (Figure 3).

**Figure 3.**
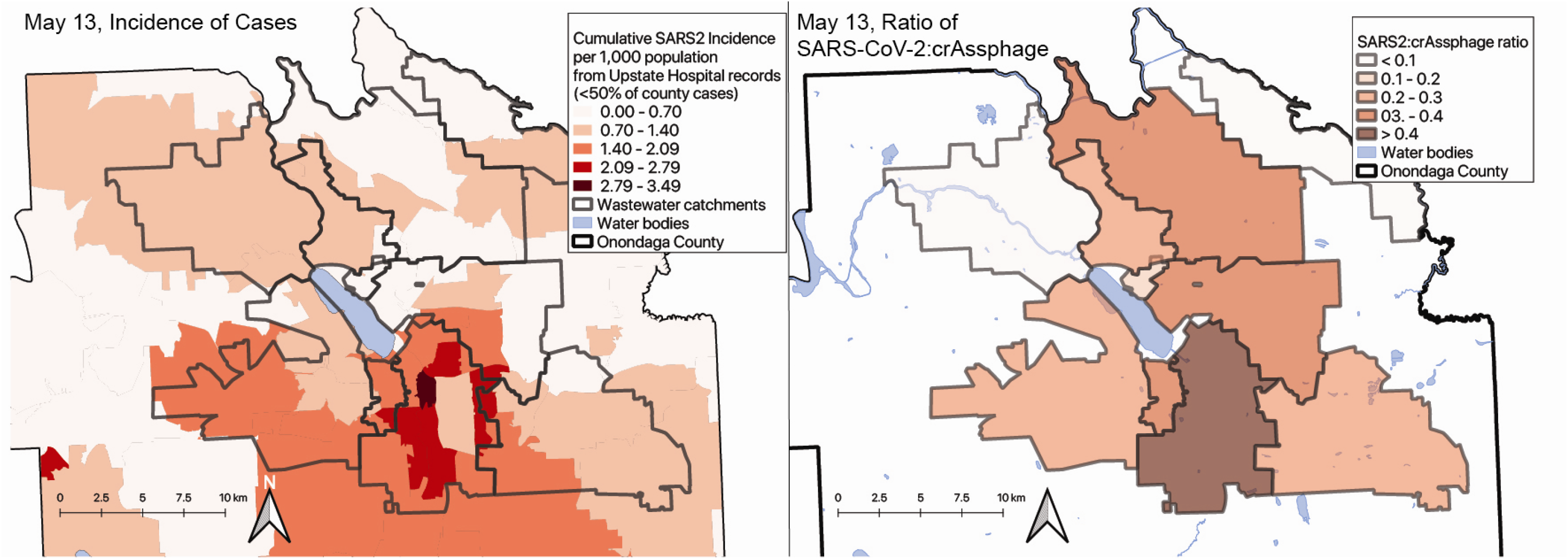
Maps of incidence of cases by zip code (left) and SARS-CoV-2:crAssphage DNA ratios (right) on May 13^th^, 2020.

## Discussion

The first demonstration that surveillance of SARS-CoV-2 in wastewater could be used to inform the public health response to COVID-19 instantly expanded the tools available to fight the pandemic^4^. However, many recent reports of SARS-CoV-2 detection in wastewater are limited by the low levels of viral RNA recovered which can limit quantitative interpretation^4, 7^. Here, we report the optimization of a simple sucrose cushion ultracentrifugation method for the recovery of SARS-CoV-2 from wastewater that provided quantitative results from a relatively small sample size (20 ml) in about 8 hours depending on the number of samples processed at one time. We understand that the requirement of an ultracentrifuge is unrealistic for many and that other approaches will be required in the absence of this equipment. Nonetheless, using a single centrifuge, we estimate a modest throughput of about 60 samples in 24 hours with these methods.

Much of the prior work on crAssphage, specifically those targeted by the CPQ_056 assay, has been focused on its utility as an indicator of human fecal pollution in natural waterbodies^11, 13–15^. Many of the same attributes that make crAssphage an attractive indicator organism, such as its prevalence and abundance in the human population as well as its scarcity in other hosts^11, 16^, are also helpful in gauging the degree of SARS-CoV-2 transmission within the community. In addition to using crAssphage as an abundant surrogate for SARS-CoV-2 during method development and optimization, crAssphage can be used to ensure sufficient viral recovery, which may become an important quality assurance measure when comparing wastewater surveillance data within and between labs. Furthermore, like SARS-CoV-2, crAssphage is subject to decay and dilution within the wastewater infrastructure and while concentrations of SARS-CoV-2 alone are difficult to interpret, the ratio of SARS-CoV-2:crAssphage is likely more robust to processes that contribute to the loss of viral nucleic acids during transport. It is conceivable that these ratios could then be used to rank catchment areas by their relative degree of transmission independent of mass-balance calculations.

Enveloped RNA viruses, like SARS-CoV-2, are known to be less resilient under environmental conditions than non-enveloped DNA viruses, like crAssphage^17^. Furthermore, DNA released from lysed viral particles is thought to be more resilient to degradation than RNA. The hypothesized rapid decay of SARS-CoV-2 compared to crAssphage would likely result in the underestimation of SARS-CoV-2 transmission within the community when using this approach. Decay rates of the two viruses and their genetic material within water infrastructure are needed to further refine predictions of transmission within the population using this approach. Decay may also play a larger role in larger service areas with longer average wastewater transit times and may explain why only low levels of SARS-CoV-2 were detected in some areas with known cases.

In summary, we demonstrated that an ultracentrifugation method using a sucrose cushion can be used for quantitative environmental surveillance of SARS-CoV-2 transmission. While a more quantitative analysis is underway, we further showed that the ratio of SARS-CoV-2 RNA:crAssphage DNA found in wastewater may be spatially associated with incidence of the disease and could potentially be used to guide public health and economic intervention strategies. Regional or national surveillance of wastewater, in conjunction with clinical testing, may provide a robust decision-making platform that authorities can use to continue restarting local economies while prioritizing public health. Furthermore, frequent and widespread wastewater surveillance has the potential to indicate when and where a resurgence of SARS-CoV-2 or outbreaks of future pathogens might occur.

## Data Availability

All data are in the process of being made publicly available.

## Acknowledgments

This work would not have been possible without seed funding from Syracuse University, the Environmental Data Science Initiative at SUNY-ESF, and the SUNY Discovery Fund. We thank Frank Mento and the sanitation engineers at Onondaga County Water Environment Protection for the collection of samples and provision of supporting data. We also thank Joshua Powell for his technical advice on ultracentrifugation and the sucrose cushion technique.

## Supplementary Material

### Sucrose Cushion Optimization

#### Concentration and Centrifugation Time Assessment

To optimize the sucrose cushion purification method, we assessed recovery of crAssphage markers with varying sucrose concentrations and ultracentrifugation times. Three sucrose concentrations were used; 20%, 50%, and 70% (in TNE buffer [20 mM Tris-HCl (pH 7.0), 100 mM NaCl, 2 mM EDTA]) with two total replicates of each treatment. Prior to cushion purification, wastewater samples were centrifuged at 2,000 x g for 25 mins to remove large particles and debris (sample clarification). Six 20 ml aliquots were then distributed to 38.5 ml ultracentrifuge tubes. To the bottom of each tube, 12 ml of a solution of sucrose in TNE was slowly added with a 10 ml serological pipette so that the sucrose formed a distinct layer below the wastewater. A pellet was generated by ultracentrifugation at 4°C at 150,000 x g for between 20 and 150 minutes depending on the sucrose concentration (Table S1). Supernatant was then decanted, and pellets were resuspended in 200 μL 1X PBS (20% treatment) or RNAlater (50% and 70% treatments). Resuspended pellets were stored at −20°C for <12 hours prior to nucleic acid extraction using the methods previously described.

A 50% sucrose cushion combined with a 90 minute ultracentrifugation spin time resulted in the highest concentration of recovered crAssphage markers and was thus used for subsequent optimization and wastewater sample processing until it was clear shorter centrifuge times resulted in approximately the same recovery of viral nucleic acids.

**Table S1.** Recovery of crAssphage DNA under different sucrose concentrations and centrifugation times.

| Sucrose Concentration | Spin Time (Minutes) | Replicate | Mean DNA Copies crAssphage /mL Initial WW Source (+/- SD) |
| --- | --- | --- | --- |
| 20% | 20 | 1 | 1.95E4 (4.40E2) |
| 20% | 20 | 2 | 2.53E4 (5.95E2) |
| 50% | 90 | 1 | 9.04E4 (3.56E3) |
| 50% | 90 | 2 | 1.27E5 (1.88E3) |
| 75% | 150 | 1 | 3.26E4 (3.41E2) |
| 75% | 150 | 2 | 7.52E4 (8.55E2) |

#### 50% Sucrose Concentration Centrifuge Time Assessment

Further optimization was performed by assessing varying spin times with a 50% sucrose cushion (Table S2). Six samples were prepared using the methods previously described, with the omission of the initial 2,000 x g clarification spin. Pellets were generated by ultracentrifugation at 4°C at 150,000 x g for 30, 45, and 75 minutes (2 replicates per centrifuge time). Pellets were examined for physical differences (Figure S1) under the assumption that darker pellets indicated a greater concentration of impurities in the sample. Pellets were resuspended in 200 PBS (1X) and stored at −20°C for <12 hours prior to nucleic acid extraction and cDNA generation using the methods previously described.

A 45 minute centrifugation time resulted in the highest concentration of recovered crAssphage cDNA markers and approximately the same amount of crAssphage DNA markers as longer spins. Due to these recovery data, and the benefit of increased processing throughput provided by a shorter spin time, wastewater samples were purified with 45 minutes of ultracentrifugation starting on May 13th.

**Table S2.**
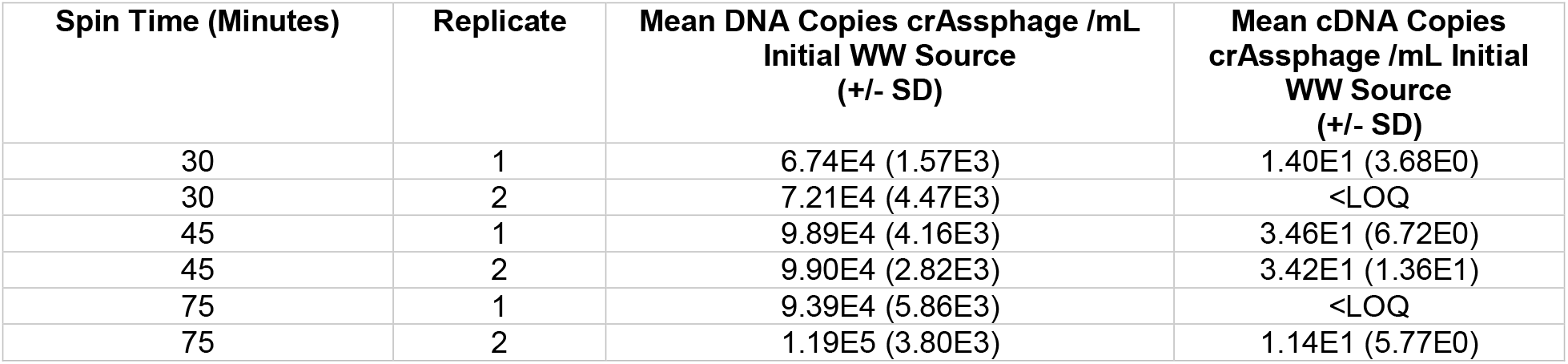
Recovery of crAssphage from the pellet after 30, 45, and 75 minute centrifugation times.

**Figure S1.**
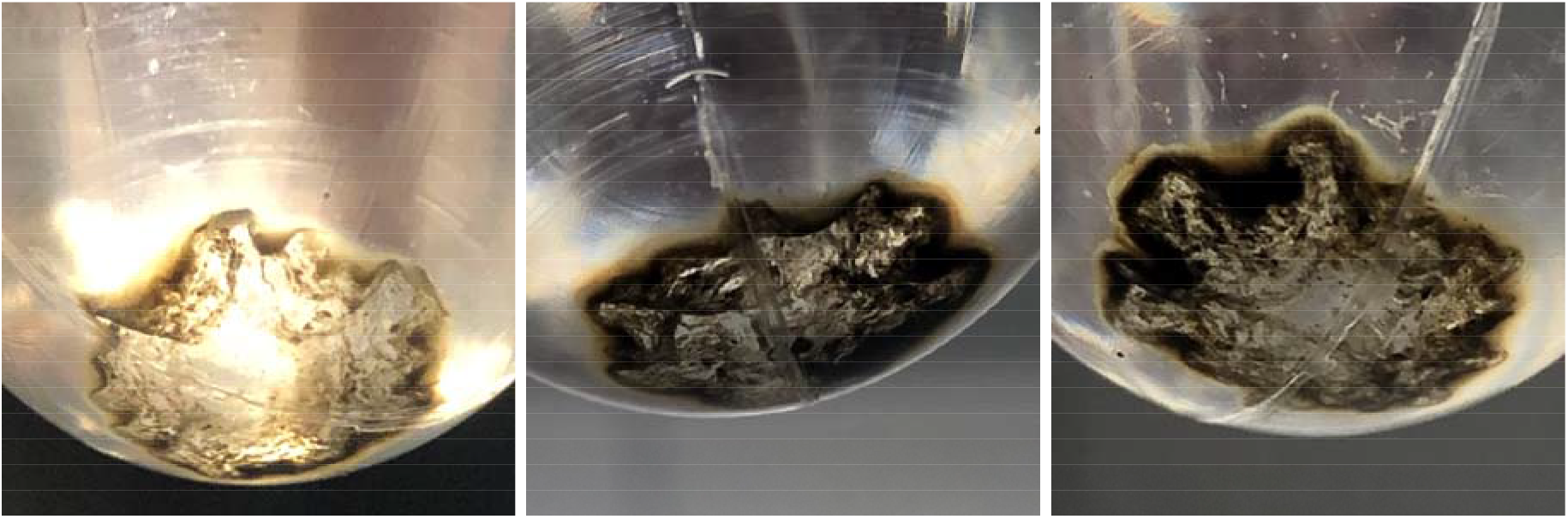
Pellets generated by 30 (left), 45 (center), and 75 (right) minutes of ultracentrifugation at 150,000 x g. Darker regions may indicate that longer spin times resulted in more impurities being pelleted.

#### Quantification of crAssphage using SYBR Green Chemistry

For optimization of our sample processing approach, SYBR Green qPCR was used to assess recovery of crAssphage markers. Reactions consisted of 12.5 μL TaqMan Environmental MasterMix, 1 μM CPQ_056 primers, 0.25 μL 10X SYBR Green dye, molecular grade water, and 2 μL template DNA for a total reaction volume of 25 μL. Thermal cycling conditions were 10 minute at 95°C, followed by 40 cycles of 95°C for 15 seconds and 60°C for 1 minute followed by a melt curve. A standard curve, consisting of purified amplicons ranging from 1×10^6^ to 5 copies/reaction, was used to convert Ct values to gene copies per reaction (Table S3).

**Table S3.** qPCR assay performance for CPQ_056 with SYBR Green detection chemistry

| Assay | Detection Chemistry | R<sup>2</sup> | Intercept | Slope | Efficiency |
| --- | --- | --- | --- | --- | --- |
| CPQ_056 | SYBR Green | 0.997 | 35.626 | -3.343 | 0.99 |

**Figure S2.**
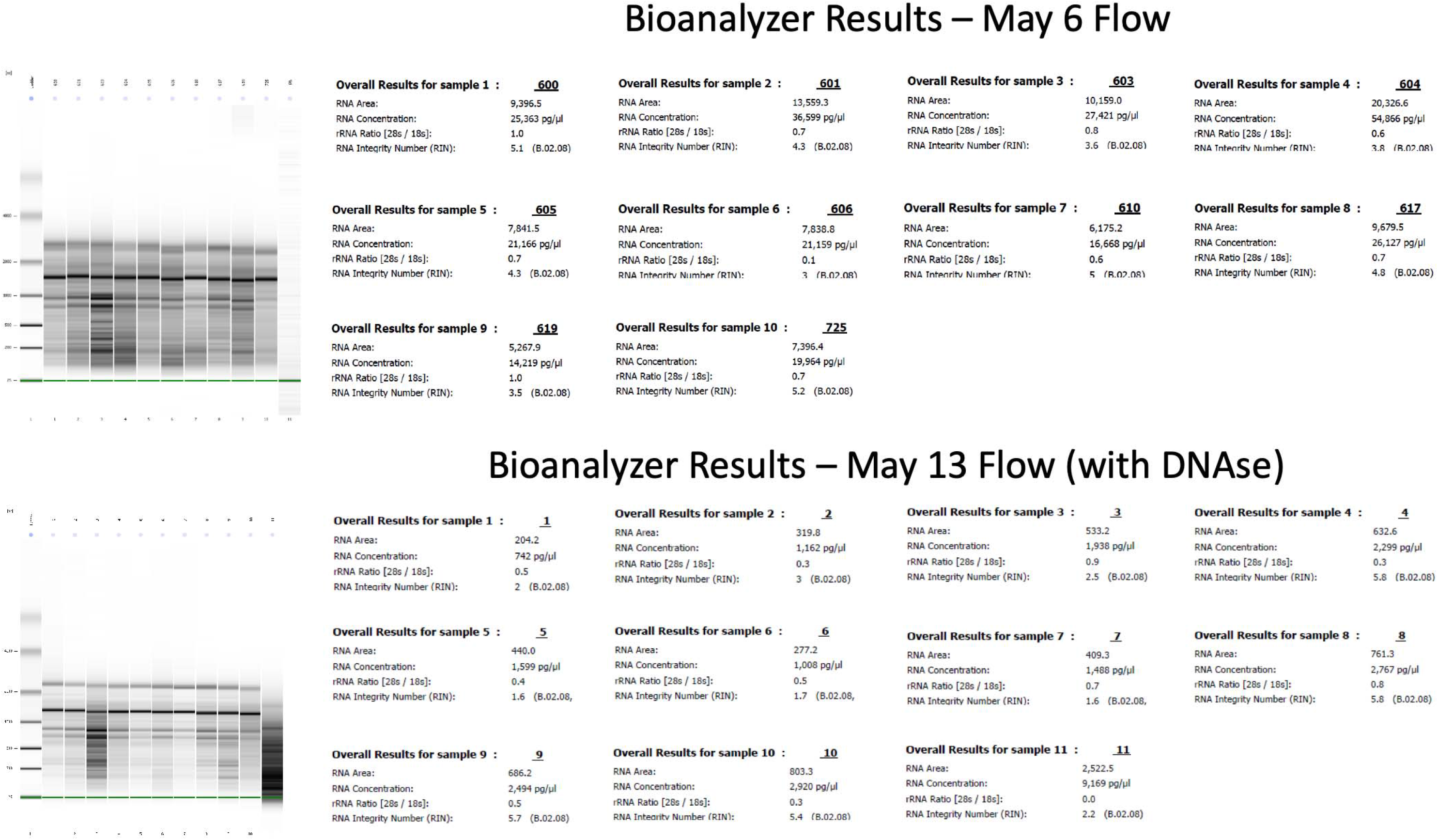
Nucleic acid quality and quantity from May 6 and May 13 samples.

## References

1. Wu, Y.; Guo, C.; Tang, L.; Hong, Z.; Zhou, J.; Dong, X.; Yin, H.; Xiao, Q.; Tang, Y.; Qu, X.; Kuang, L.; Fang, X.; Mishra, N.; Lu, J.; Shan, H.; Jiang, G.; Huang, X., Prolonged presence of SARS-CoV-2 viral RNA in faecal samples. The Lancet Gastroenterology & Hepatology 2020, 5, (5), 434–435.

2. Xing, Y. H.; Ni, W.; Wu, Q.; Li, W. J.; Li, G. J.; Wang, W. D.; Tong, J. N.; Song, X. F.; Wing-Kin Wong, G.; Xing, Q. S., Prolonged viral shedding in feces of pediatric patients with coronavirus disease 2019. J Microbiol Immunol Infect 2020.

3. Wölfel, R.; Corman, V. M.; Guggemos, W.; Seilmaier, M.; Zange, S.; Müller, M. A.; Niemeyer, D.; Jones, T. C.; Vollmar, P.; Rothe, C.; Hoelscher, M.; Bleicker, T.; Brünink, S.; Schneider, J.; Ehmann, R.; Zwirglmaier, K.; Drosten, C.; Wendtner, C., Virological assessment of hospitalized patients with COVID-2019. Nature 2020.

4. Medema, G.; Heijnen, L.; Elsinga, G.; Italiaander, R.; Brouwer, A., Presence of SARS-Coronavirus-2 in sewage. *medRxiv* 2020, 2020.03.29.20045880.

5. Wurtzer, S.; Marechal, V.; Mouchel, J.-M.; Maday, Y.; Teyssou, R.; Richard, E.; Almayrac, J. L.; Moulin, L., Evaluation of lockdown impact on SARS-CoV-2 dynamics through viral genome quantification in Paris wastewaters. *medRxiv* 2020, 2020.04.12.20062679.

6. Larsen, D.; Dinero, R. E.; Asiago-Reddy, E.; Green, H.; Lane, S.; Shaw, A.; Zeng, T.; Kmush, B., A review of infectious disease surveillance to inform public health action against the novel coronavirus SARS-CoV-2. Journal of Infectious Disease 2020.

7. Ahmed, W.; Angel, N.; Edson, J.; Bibby, K.; Bivins, A.; O’Brien, J. W.; Choi, P. M.; Kitajima, M.; Simpson, S. L.; Li, J.; Tscharke, B.; Verhagen, R.; Smith, W. J. M.; Zaugg, J.; Dierens, L.; Hugenholtz, P.; Thomas, K. V.; Mueller, J. F., First confirmed detection of SARS-CoV-2 in untreated wastewater in Australia: A proof of concept for the wastewater surveillance of COVID-19 in the community. Science of The Total Environment 2020, 728, 138764.

8. Wu, F.; Xiao, A.; Zhang, J.; Gu, X.; Lee, W. L.; Kauffman, K.; Hanage, W.; Matus, M.; Ghaeli, N.; Endo, N.; Duvallet, C.; Moniz, K.; Erickson, T.; Chai, P.; Thompson, J.; Alm, E., SARS-CoV-2 titers in wastewater are higher than expected from clinically confirmed cases. *medRxiv* 2020, 2020.04.05.20051540.

9. Randazzo, W.; Cuevas-Ferrando, E.; Sanjuan, R.; Domingo-Calap, P.; Sanchez, G., Metropolitan Wastewater Analysis for COVID-19 Epidemiological Surveillance. *medRxiv* 2020, 2020.04.23.20076679.

10. Farkas, K.; Cooper, D. M.; McDonald, J. E.; Malham, S. K.; de Rougemont, A.; Jones, D. L., Seasonal and spatial dynamics of enteric viruses in wastewater and in riverine and estuarine receiving waters. Science of The Total Environment 2018, 634, 1174–1183.

11. Stachler, E.; Kelty, C.; Sivaganesan, M.; Li, X.; Bibby, K.; Shanks, O. C., Quantitative CrAssphage PCR Assays for Human Fecal Pollution Measurement. Environmental science & technology 2017, 51, (16), 9146–9154.

12. Institut Pasteur, Real-time RT-PCR assays for the detection of SARS-CoV-2. 2020.

13. Ahmed, W.; Gyawali, P.; Feng, S.; McLellan, S. L., Host Specificity and Sensitivity of Established and Novel Sewage-Associated Marker Genes in Human and Nonhuman Fecal Samples. Applied and environmental microbiology 2019, 85, (14), e00641–19.

14. Farkas, K.; Adriaenssens, E. M.; Walker, D. I.; McDonald, J. E.; Malham, S. K.; Jones, D. L., Critical Evaluation of CrAssphage as a Molecular Marker for Human-Derived Wastewater Contamination in the Aquatic Environment. Food Environ Virol 2019, 11, (2), 113–119.

15. Stachler, E.; Bibby, K., Metagenomic Evaluation of the Highly Abundant Human Gut Bacteriophage CrAssphage for Source Tracking of Human Fecal Pollution. Environmental Science & Technology Letters 2014, 1, (10), 405–409.

16. Dutilh, B. E.; Cassman, N.; McNair, K.; Sanchez, S. E.; Silva, G. G. Z.; Boling, L.; Barr, J. J.; Speth, D. R.; Seguritan, V.; Aziz, R. K.; Felts, B.; Dinsdale, E. A.; Mokili, J. L.; Edwards, R. A., A highly abundant bacteriophage discovered in the unknown sequences of human faecal metagenomes. Nature Communications 2014, 5, (1), 4498.

17. Ye, Y.; Ellenberg, R. M.; Graham, K. E.; Wigginton, K. R., Survivability, Partitioning, and Recovery of Enveloped Viruses in Untreated Municipal Wastewater. Environmental science & technology 2016, 50, (10), 5077–5085.

